# Keyphrase Identification Using Minimal Labeled Data with Hierarchical Contexts and Transfer Learning

**DOI:** 10.1101/2023.01.26.23285060

**Authors:** Rohan Goli, Keerthana Komatineni, Shailesh Alluri, Nina Hubig, Hua Min, Yang Gong, Dean F. Sittig, Lior Rennert, David Robinson, Paul Biondich, Adam Wright, Christian G. Nøhr, Timothy D. Law, Arild Faxvaag, Aneesa Weaver, Ronald W. Gimbel, Xia Jing

**Author notes:** Correspondence Author: Xia Jing.

## Abstract

**Background:** Interoperable clinical decision support system (CDSS) rules provide a pathway to interoperability, a well-recognized challenge in health information technology. Building an ontology facilitates creating interoperable CDSS rules, and identifying the keyphrases (KP) from the existing literature can be a first step for building an ontology. Ontology construction requires curation by human domain experts (HDE) traditionally, and modern natural language processing (NLP) techniques can be a critical complementary component nevertheless requires human proficiency, consensus, and contextual understanding for data labeling.

**Methods:** We present a semi-supervised KP identification framework using a hierarchical-attention BiLSTM-CRF (Hier-Attn-BiLSTM-CRF) with word-, sentence-, and document-level attention. A domain-adapted sciSpacy model was used to generate synthetic labels for bootstrap training, followed by fine-tuning with minimal HDE-labeled data. We then evaluated robustness through component ablation, comparison with fine-tuned biomedical transformers (BioBERT, SciBERT, PubMedBERT) with bootstrap confidence intervals, train-split sensitivity, multi-seed variance analysis, error analysis, and benchmarking against public corpora (KPBioMed, PubMedAKE).

**Results:** The Hier-Attn-BiLSTM-CRF is competitive with fine-tuned transformer baselines on the HDE labeled dataset (GS42: ∼44 vs. ∼46 F1; GS91: 61.3 vs. ∼54 F1), through an explicit hierarchical inductive bias over word-, sentence-, and document-level representations, complementing the implicit contextual modeling of the transformer. Controlled ablation identifies gold-standard fine-tuning as the dominant performance lever. Mixing HDE and synthetic labels in 2:100–4:100 ratios improved performance without exhausting the human-labeled set too quickly. Models trained on sparser public corpora transferred poorly to CDSS across all architectures, underscoring the value of in-domain synthetic labels.

**Conclusions:** This feasibility study demonstrates a practical, resource-efficient framework for CDSS KP identification under limited HDE annotation. The contribution lies in integrating established components—domain-adapted synthetic labels, hierarchical attention, and minimal gold-standard fine-tuning—for the CDSS sub-domain. A full downstream evaluation of the role of the NLP pipeline for CDSS ontology construction is the primary next step.

The code is available on GitHub: https://github.com/xjing16/cdss4pcp_nlpml_pipeline.

## 1. Background

Interoperability [1, 2] is a well-recognized challenge in health informatics. Clinical decision support systems (CDSSs), especially rule-based ones, have been effective in improving healthcare quality [3, 4], but developing, maintaining, and sharing rules among institutions remains resource-demanding. A CDSS ontology [5, 6] that uses unambiguous concepts and their relationships can facilitate the interoperability of such rules [4, 7].

In a text corpus, concepts are identified as ordered sequences of words (N-grams), namely keyphrases (KP). Human domain expert (HDE)-identified KPs constitute the gold standards (GSs) that seed ontology construction after HDE panel review and consensus [7]. Automating candidate KP identification through NLP can complement this manual process [8, 9].

### 1.1. Related work

KP identification can be approached through statistical and unsupervised methods such as TF-IDF [10, 11], BM25 [12], and graph-based ranking algorithms (PageRank [13], MultiPartiteRank [14], PositionRank [15], TopicRank [16]), though these methods do not consider document-level context. Supervised methods—Naïve Bayes [17], Decision Trees [18], SVM [19]—treat KP identification as binary classification but cannot capture the sequential dependencies of N-gram phrases. The BiLSTM-CRF [20] model frames it as a sequence tagging problem with superior performance [21] but does not address limited HDE-labeled data.

Named entity recognition (NER) [22] extracts N-gram entities into predefined categories (e.g., drug, gene, disease) but cannot identify domain-specific concepts or topics. NER and KP identification are complementary objectives that differ in scope (Figure 1).

**Figure 1.**
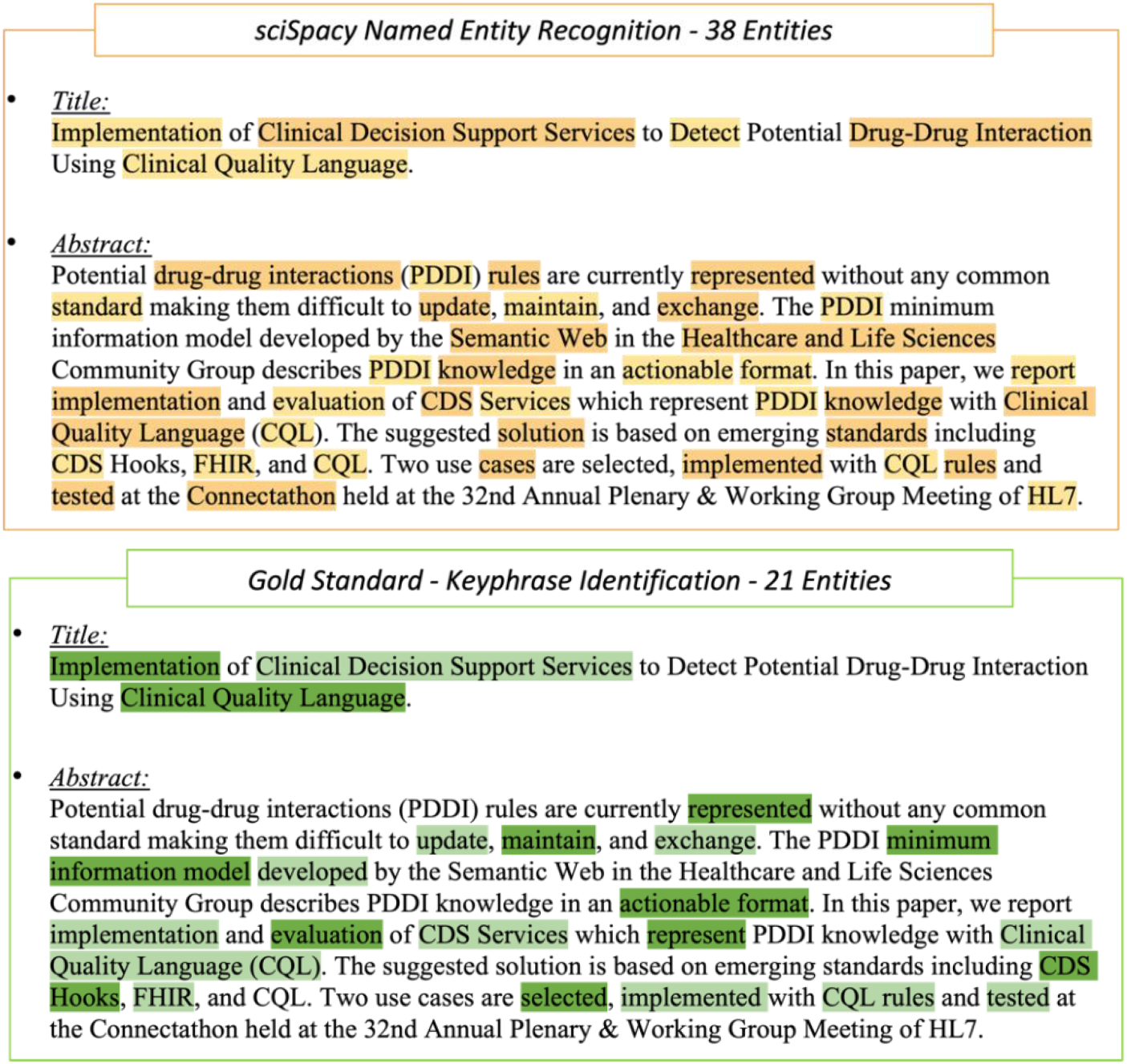
Entities identified on a sample CDSS abstract demonstrate differences between human-labeled Gold Standards and NER by sciSpacy.

Deep learning approaches such as BERT [23-25] outperform on many NLP tasks but require significant computational resources and labeled data for fine-tuning. Lightweight encoder-decoder models [26, 27] using LSTM-based encoders [28] and CRF-based decoders [29] offer a resource-efficient alternative. Bidirectional attention for LSTM enhances KP prediction [30], and our approach builds on the architectures of Yang et al. and Xu et al. [31, 32] by augmenting a document-level attention layer.

### 1.2. Domain Adaptation and Language Modeling

Domain adaptation [33, 34] transfers knowledge from pre-trained models to related domains, reducing the need for ground-up training. sciSpacy [35], a domain adaptation of spaCy [36] trained on biomedical corpora, provides the foundation for our synthetic label generation. Language models (LM) [37] estimate word distributions using bidirectional context [38], enabling domain-specific knowledge transfer [39].

### 1.3. Our Contribution

We contribute a CDSS-specific integration of established components: (1) a keyphrase task formulation targeting CDSS ontology-relevant phrases; (2) a synthetic-label workflow built on domain-adapted sciSpacy; (3) a document-level attention layer added to the BiLSTM-CRF; and (4) a gold-standard fine-tuning step that achieves clean-label performance with minimal HDE labels. Our main objectives are: identifying KPs with long-range contextual dependencies through hierarchical attention; creating high-quality synthetic labels via domain adaptation; and optimizing the fine-tuning process with limited HDE-labeled data.

We designed a pipeline comprising a Hier-Attn-BiLSTM encoder and a CRF decoder that learns N-gram entity patterns using hierarchical attention at word, sentence, and document levels [30, 31].

## 2. Methods

We designed a pipeline comprising a Hier-Attn-BiLSTM encoder and a CRF decoder that learns N-gram entity patterns using hierarchical attention at word, sentence, and document levels [30, 31].

### 2.1. Task Formulation

KP identification is formulated as a sequence labeling task: for a document with *m* sentences, each containing *n* tokens, the model outputs a BIO tag sequence [40] where B-KP marks the first token of a KP, I-KP marks continuation tokens, and O marks non-KP tokens (Figure 2).

**Figure 2.**
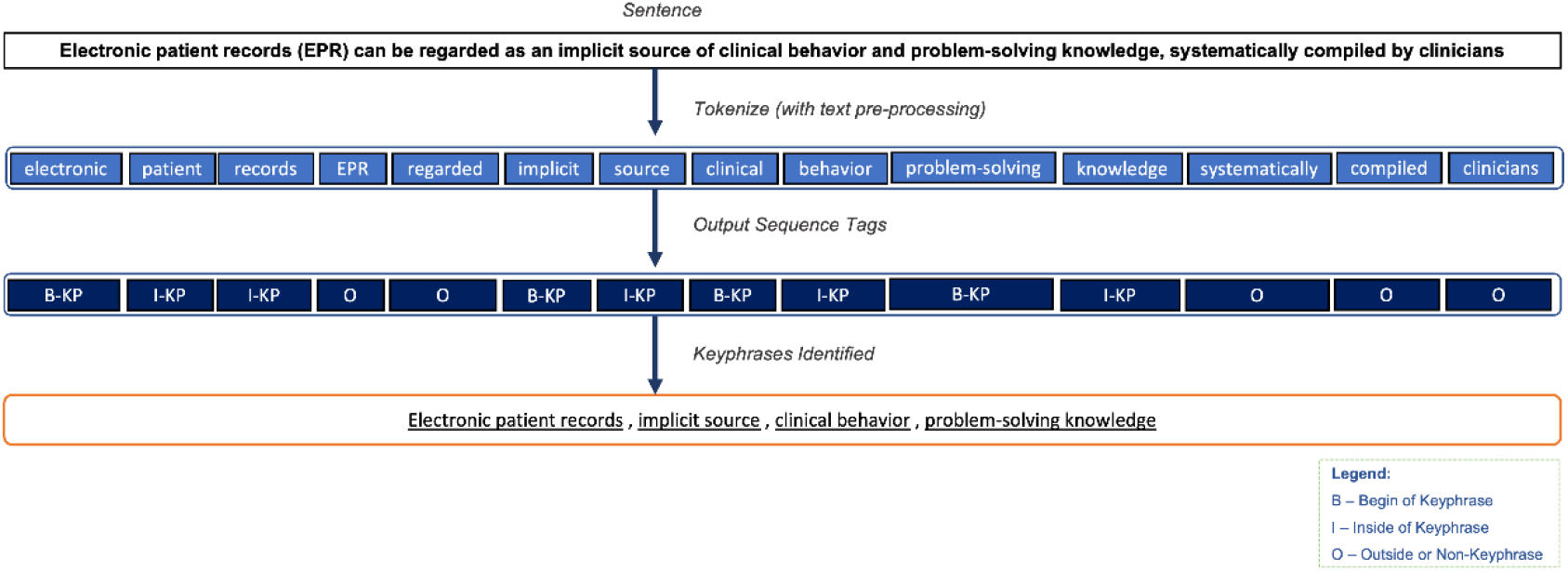
Flow of labeling Keyphrases (KP) from a sentence.

In BIO token tagging [40], the first N-gram word is labeled B-KP, the rest are labeled I-KP, and the non-KP tokens are marked as O. Figure 2 presents a BIO token tagging example with an input document and the sequence tag (B-KP/I-KP/O) output, where the keyphrases can be generated by decoding the output tags.

### 2.2. Pipeline Overview

As shown in Figure 3, the pipeline proceeds as follows: (1) generate synthetic labels using a domain-adapted sciSpacy [35] model [32, 41]; (2) pre-train Word2Vec word embedding (WE) [42] and bidirectional language (BiLM) models on the unlabeled corpus; (3) transfer pre-trained weights into the Hier-Attn-BiLSTM layers; (4) train the model using synthetic labels by feeding one document at a time, applying hierarchical attention (Appendix A) and CRF decoding; and (5) fine-tune with minimal HDE-labeled data.

**Figure 3.**
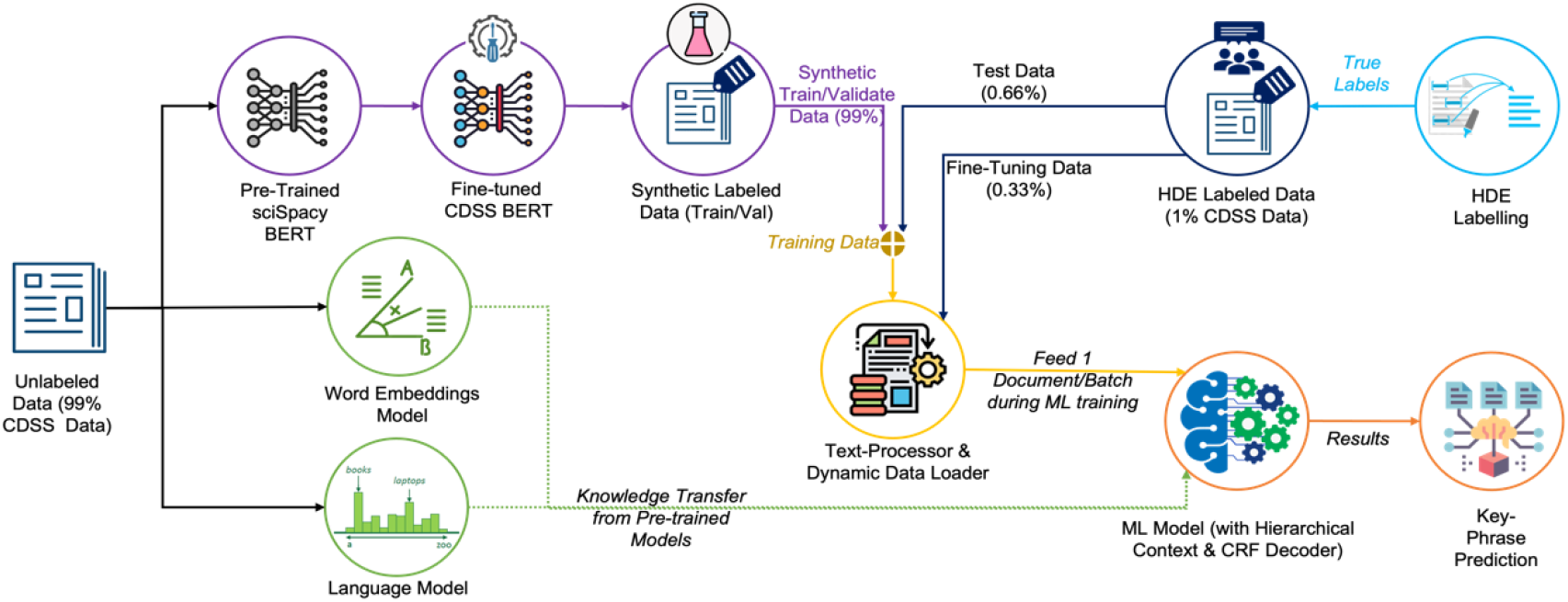
High-level design of the proposed NLP pipeline for KP identification.

### 2.3. Synthetic labels

We performed domain adaptation of sciSpacy [35] on the CDSS dataset to generate intermediate synthetic KPs, then fine-tuned the model to produce the final synthetic labels in BIO format. These synthetic labels bootstrap the training process (Figure 3) in the absence of large-scale HDE annotated labels.

### 2.4. Pre-training

We pre-trained a Word2Vec [42] embedding model (300-dimensional) on the CDSS corpus. A shallow bidirectional RNN [43] language model (BiLM) learns word distributions by computing the conditional probability of each word based on forward and backward context [37], with each direction encoded into fixed-dimensional vectors via a soft-max layer. Transfer is a direct weight transfer: the pre-trained BiLM’s recurrent LSTM parameters (weights and biases) matching the encoder’s first BiLSTM layer in name and shape are copied in as initialization, while the soft-max head, attention layers, and CRF decoder are randomly initialized. It is neither embeddings-only nor a hidden-state initialization, and the copied weights are updated during training.

### 2.5. Hierarchical-Attention-BiLSTM-CRF Model

#### 2.5.1. Encoder

This architecture is adopted from Zichao Yang, Guohai Xu, and Luo L et al. (Figure 4) [31, 32, 44]. To capture the context of the KP, we encode one document at-a-time to capture document-level context with a stacked BiLSTM [45] whose embedding matrix is initialized from pre-trained Word2Vec weights and whose first BiLSTM layer is initialized from the shape-matched recurrent parameters of the pre-trained BiLM [32]. We used all the sentences in a document, *d* = (*s*_1_, *s*_2_, …, *s*_*m*_), where each sentence is represented by *s*_*i*_ = (*w*_*i*1_, *w*_*i*2_, …, *w*_*in*_) and its words by *w*_*it*_ ∀*t* ∈ [1, *n*]. We embed the words into a vector (*x*_*it*_) through an *embedding matrix* (*W*_*e*_). BiLSTM summarizes the bidirectional context information where each word vector’s hidden state (*h*_*it*_) is obtained by concatenating the forward 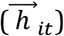 and backward 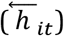 hidden state vectors, i.e.,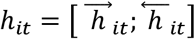. The hidden state vector provides sentence-level context to each word [26].

**Figure 4.**
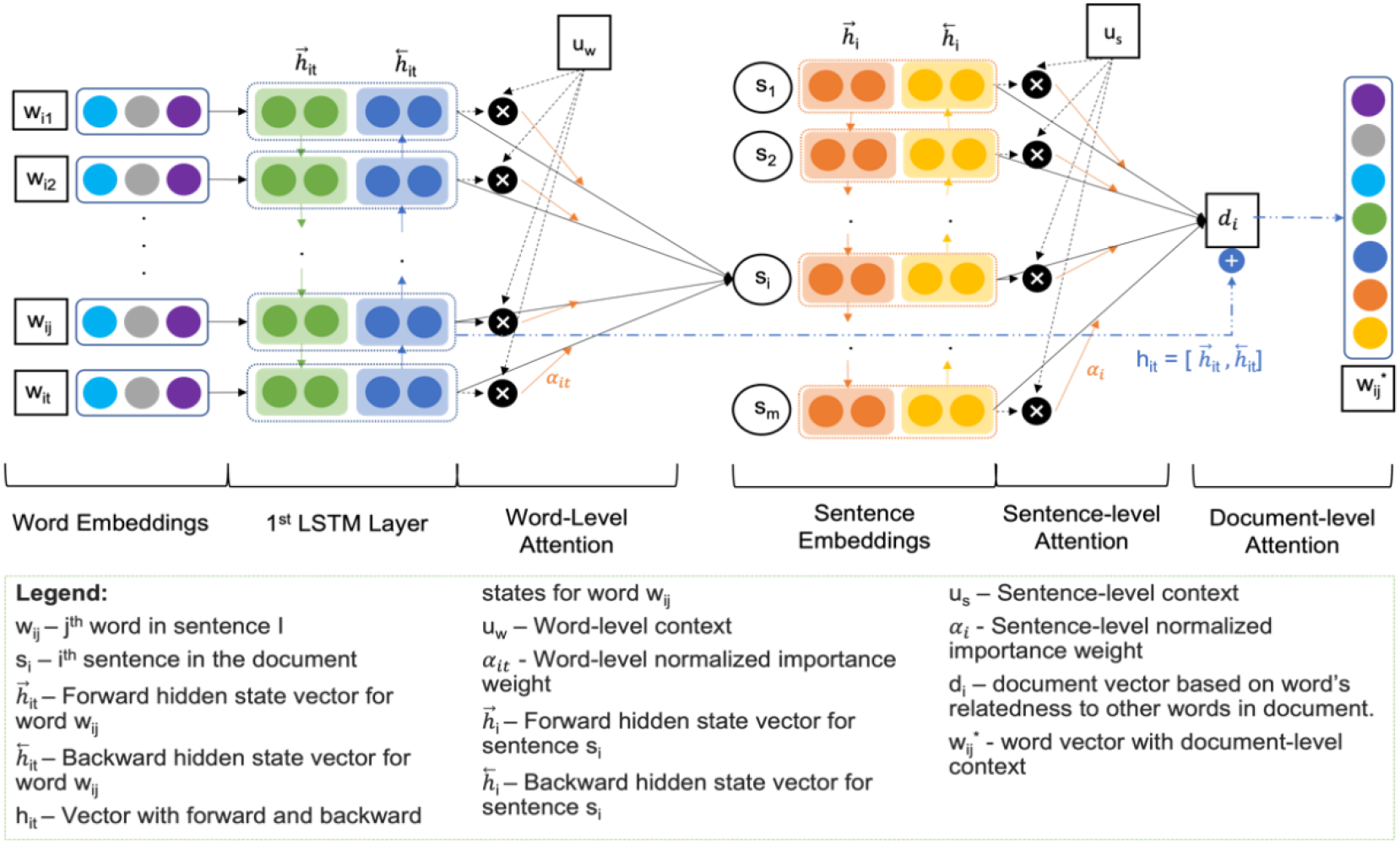
Detailed encoder - word encoding with Hierarchical-Attention-BiLSTM with document-level context.

We calculate word similarity (*u*_*it*_) using a neural network’s parameter for weighted matrix (*W*_*w*_) and word representation (*h*_*it*_) given by BiLSTM along with bias (*b*_*w*_) [46] and the **word-level attention** by aggregating the *h*_*it*_ and *u*_*it*_ using a word-level context vector (*u*_*w*_) [10, 11] to get a word-level normalized importance weight (*α*_*it*_). Finally, we compute the sentence vector (*s*_*i*_) as a weighted sum of word representations as shown in Eq. (A. 1)(A. 2)(A 3). Initially, *u*_*w*_ is the neural network parameter with random initialization, learned during the training process.

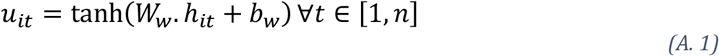

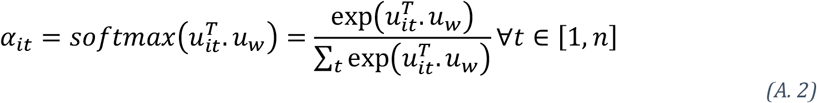

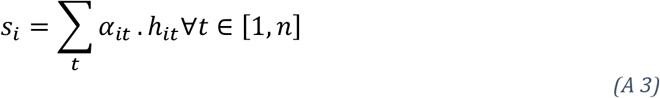

Similarly, a document vector can be computed using **sentence-level attention** over the sentence vectors (*s*_*i*_) [10, 11] and concatenating the forward 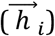 and backward 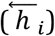 states to encode a sentence, 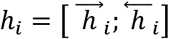 based on neighbor sentences.

To estimate the sentence-level context vector (*u*_*s*_, *B*. 1, *B*. 2, *B*. 3), first, we used neural network parameter for weighted matrix (*W*_*s*_), sentence representation (*h*_*i*_) and bias (*b*_*s*_) to calculate sentence similarity (*u*_*i*_). Second, we randomly initialize *u*_*s*_, to calculate the sentence-level normalized importance weight (*α*_*i*_), which yields a **document vector(***d*_*i*_) for each word representing the sentences that are important to consider for a given word while identifying it as a KP as provided [31].

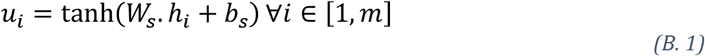

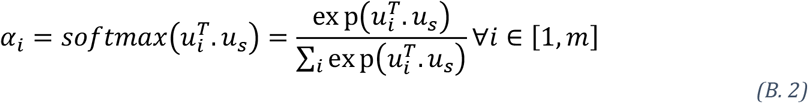

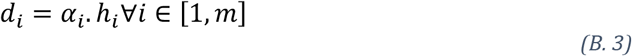

Unlike the previous work proposed by Guohai Xu et al. [32], we concatenated the first LSTM’s hidden local state (*h*_*it*_) with the document vector (*d*_*i*_) into a new vector [*h*_*it*_ ; *d*_*i*_]∀*t* ∈ [1, *n*], given the word’s relatedness to other words in the document. That is, providing document-level context to each word. Next, the extended representation further used to identify the labels.

#### 2.5.2. Decoder

Following Ling Luo et al. [44], we use a CRF [29] decoder that produces confidence scores for each label (B-KP/ I-KP/ O). The sentence score is the sum of transition and network scores Eq.(C. 1), where the tagging transformation matrix(T) is learned during training. The conditional probability of a tag path is computed via soft-max normalization Eq. (C. 2), and Viterbi decoding [47] recovers the optimal sequence Eq. (C. 3).

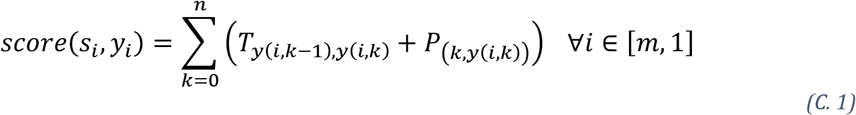

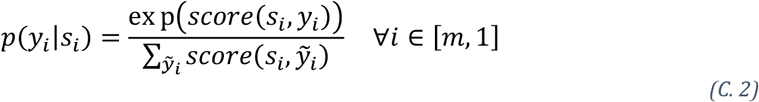

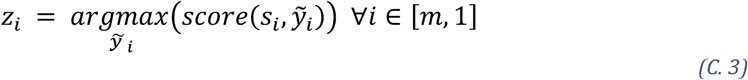

## 3. Experiments and results

This section presents the dataset (4.1), synthetic label creation (4.2), preparation steps (4.3), model improvement strategies (4.4) and additional robustness experiments (4.5). Unless noted, reported values are strict entity-level SeqEval micro-F1 (%) [48].

### 3.1. Dataset

The text corpus was obtained from PubMed by retrieving the CDSS (as the Medical Subject Headings, MeSH) literature. The articles with a valid PubMed Identifier (PMID) were selected. The corpus profile presents in Tables 1 and 2. Appendix B provides detailed information on the dataset at the various stages during text preprocessing.

**Table 1.** The profile of the CDSS corpus.

|  | **FC** | **FC with PMIDs** | **GS (+8 ACM)** | **No/little abs.** | **Final/total dataset** |
| --- | --- | --- | --- | --- | --- |
| Articles | 3545 | 3326 | 133 | 99 | 3281 |

FC – Full CDSS dataset
No/little abs. - Abstracts having fewer than 3 sentences including the title.

**Table 2.** The CDSS Datasets for training, validation, and testing.

|  | **Total dataset** | **Synthetic KP labeled dataset** | **Training** | **Validation** | **Testing (GS91)\*** | **GS(GS42)\*** |
| --- | --- | --- | --- | --- | --- | --- |
| Articles | 3281 | 3148 | 1049 | 2099 | 91 | 42 |

\* GS91 and GS42 are 2 sets of HDE-labeled datasets.

Of the total dataset retrieved from PubMed (3545 abstracts): 3326 abstracts were kept after XML parsing, and 133 of them were labeled by HDE (Table 1, Appendix B). During preprocessing, we removed articles with abstracts of 3 or fewer sentences, treating the title as a single sentence.

HDE labeled datasets are grouped into 2 sets (GS91 and GS42). GS91 was unseen data to test the model’s performance, and the GS42 was used to fine-tune the ML model. Cohen’s kappa rates for the first 42 (GS42) abstracts were 0.93 (annotators 1 and 2) and 0.73 (annotators 1 and 3) [49]. For the second set of abstracts (GS91), Cohen’s kappa rates were 0.87 (annotators 1 and 2) and 0.97 (annotators 1 and 3). For the remaining 3,148 articles, they were labeled by the synthetic KP, which were split into training and validation data sets in 1:2 ratio (Table 2).

Additionally, we used KPBioMed [50] and PubMedAKE [51] public datasets as baselines to understand and evaluate the use of synthetic labels in the training process. For comparison, we formulated label-sparsity ratio (fraction of tokens that belong to KP in a document) where *ρ*_*public*_ ≈ 0.06 − 0.08 ≪ *ρ*_*GS*_ ≈ 0.19 − 0.20. The public datasets are 3-5x sparser in KP density than the Gold-Standard and Synthetic datasets (Table 3).

**Table 3.** Different Corpora Statistics.

| **Corpus** | **Role** | **Sample Size** | $\rho = \frac{\#\{KP\ Tokens\}}{\#\{Tokens\}}$ | **KP Style** | **Dataset Size** | **Notes** |
| --- | --- | --- | --- | --- | --- | --- |
| Synthetic CDSS | In-Domain | 1049 | 0.261 | Dense weak labels | 3148 | Synthetic KP labelled CDSS set |
| KPBioMed | Public | 90000 | 0.055 | Sparse author KP | 5.6 M total | Large Dataset, Author Assigned, ~1.8% sampled, sparsest corpus |
| PubMedAKE | Public | 73810 | 0.085 | Sparse author KP | 843,269 total | Author Assigned, 10% sampled, |
| GS42/GS91 | HDE Labels | 42 / 91 | 0.188 / 0.203 | Dense expert annotation | 133 | CDSS Expert (HDE) Gold Standard |

\* GS91 and GS42 are 2 HDE-labeled datasets.

### 3.2. Synthetic label creation

To evaluate the quality of the created synthetic labels, we experimented with different unsupervised algorithms (namely, PositionRank, MultiPartiteRank, and TopicRank)[10-16] and NER (i.e., sciSpacy). BERT-based sciSpacy [35] outperformed other unsupervised methods (Table 4).

**Table 4.** Evaluation of synthetic KP generated with different approaches.

| Approach | Accuracy | Misclassification | Precision | Recall | Specificity | F1-Score |
| --- | --- | --- | --- | --- | --- | --- |
| sciSpacy | 0.69 | 0.31 | 0.36 | **0.81** | 0.66 | **0.50** |
| PositionRank | 0.76 | 0.23 | **0.39** | 0.36 | 0.86 | 0.38 |
| MultiPartiteRank | 0.76 | 0.24 | 0.38 | 0.36 | 0.86 | 0.37 |
| TopicRank | 0.77 | 0.23 | **0.39** | 0.36 | **0.87** | 0.37 |

### 3.3. Preparation of KP identification model

Domain adaptation of sciSpacy improved synthetic label quality substantially (Appendix D, Table D1; Figure 5). BIO token tagging slightly outperformed BILOU (Appendix D, Table D2), and stemming degraded performance (Appendix D, Tables D3– D4), so we used BIO encoding with non-stemmed text.

**Figure 5.**
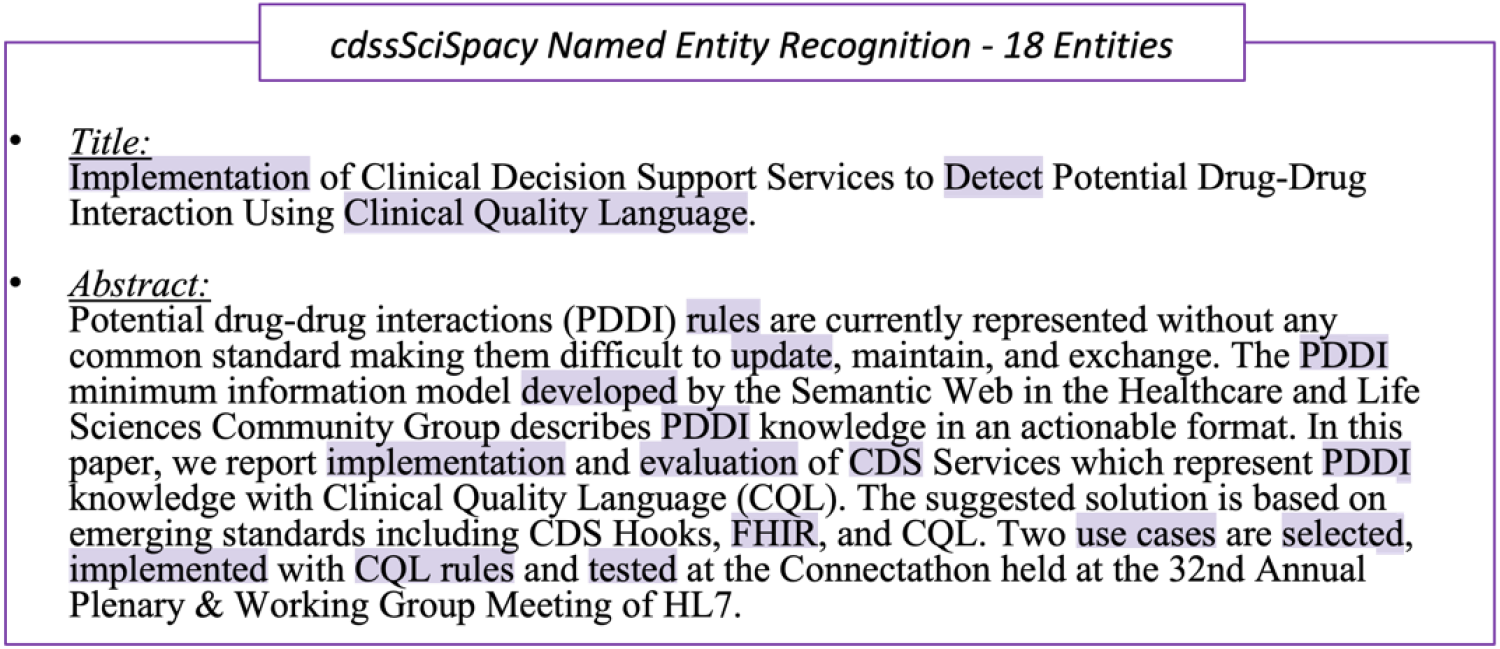
Entities identified on sample CDSS article context with CDSS-domain adapted sciSpacy NER.

The Hier-Attn-BiLSTM-CRF was trained for 30 epochs with: WE dimension 300, LSTM hidden dimension 256, dropout 0.2, batch size 1 (one document at a time for document-level context), max sentence length 128, Word2Vec embeddings, 1:2 train-validation split, and sciSpacy en_core_sci_lg for synthetic label generation.

### 3.4. KP identification ML model improvement

We compared encoding combinations including character-level embeddings, CNN-based text features (POS tag, TF-IDF, Position of First Occurrence [52]), and transformer encoders (BioBERT [53], PubMedBERT [54], sciBERT [55]) against our Hier-Attn-BiLSTM-CRF (Table 5, Figures 6–7). The hierarchical model achieves the highest precision (0.75 synthetic, 0.60/0.61 GS42/GS91) and accuracy (up to 0.92) across all models, matching transformers on GS42 and outperforming them on GS91 while maintaining a lightweight footprint.

**Table 5.** Comparison of evaluations on different contextual levels of attention.

| Model | Encoder Details | Runs | Train Dataset | Test Dataset | Precision | Recall | Accuracy | F1-Score |
| --- | --- | --- | --- | --- | --- | --- | --- | --- |
| BiLSTM-CRF | BiLSTM (+WE) | 1 | 1049 Synthetic | 2099 Synthetic | 0.72 | 0.66 | **0.92** | 0.69 |
|  |  |  |  | 42 GS | 0.54 | 0.46 | 0.86 | 0.49 |
|  |  |  |  | 91 GS | 0.59 | 0.48 | **0.88** | 0.53 |
| BiLSTM-CRF | BiLSTM (+WE + CE) | 1 | 1049 Synthetic | 2099 Synthetic | 0.70 | 0.70 | 0.85 | 0.70 |
|  |  |  |  | 42 GS | 0.52 | 0.56 | 0.78 | 0.54 |
|  |  |  |  | 91 GS | 0.58 | 0.53 | 0.77 | **0.55** |
| BiLSTM-CRF | BiLSTM (+WE + CE) + CNN (Text Features) | 1 | 1049 Synthetic | 2099 Synthetic | 0.73 | **0.71** | 0.85 | **0.72** |
|  |  |  |  | 42 GS | 0.56 | 0.55 | 0.78 | 0.55 |
|  |  |  |  | 91 GS | 0.58 | 0.55 | 0.78 | 0.57 |
| **Hier-Attn-BiLSTM-CRF (our method)** | BiLSTM (+WE) + Hierarchical Context (Word-level sentence-level attentions) | 1 | 1049 Synthetic | 2099 Synthetic | **0.75** | 0.68 | **0.92** | **0.71** |
|  |  |  |  | 42 GS | **0.6** | 0.5 | **0.88** | 0.54 |
|  |  |  |  | 91 GS | **0.61** | 0.5 | **0.88** | **0.55** |
| BioBERT | Bidirectional Encoder Representations from Transformers | 1 | 1049 Synthetic | 2099 Synthetic | 0.55 | 0.50 | 0.81 | 0.52 |
|  |  |  |  | 42 GS | 0.39 | 0.54 | 0.80 | 0.45 |
|  |  |  |  | 91 GS | 0.51 | 0.56 | 0.83 | 0.54 |
| PubMedBERT |  | 1 | 1049 Synthetic | 2099 Synthetic | 0.56 | 0.50 | 0.81 | 0.53 |
|  |  |  |  | 42 GS | 0.39 | 0.58 | 0.80 | 0.47 |
|  |  |  |  | 91 GS | 0.50 | 0.60 | 0.83 | 0.54 |
| sciBERT |  | 1 | 1049 Synthetic | 2099 Synthetic | 0.58 | 0.53 | 0.82 | 0.56 |
|  |  |  |  | 42 GS | 0.38 | **0.59** | 0.80 | 0.46 |
|  |  |  |  | 91 GS | 0.48 | **0.62** | 0.82 | 0.54 |

**Figure 6.**
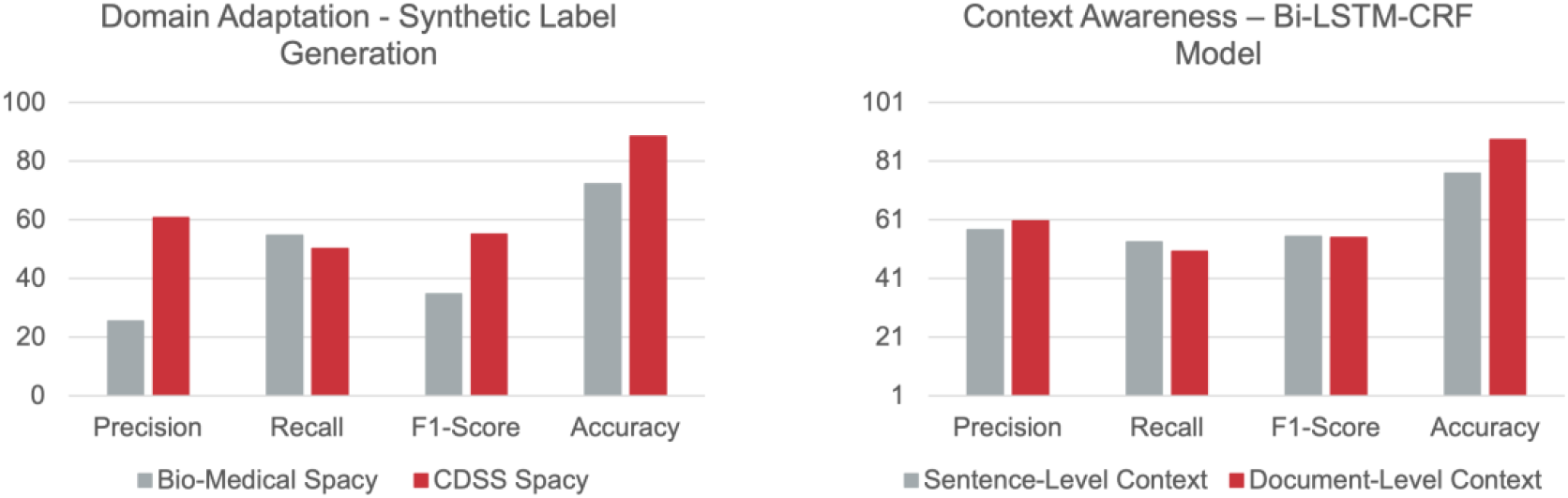
Comparison of results for domain adaptation and hierarchical context (document context through word-level and sentence-level attention).

**Figure 7.**
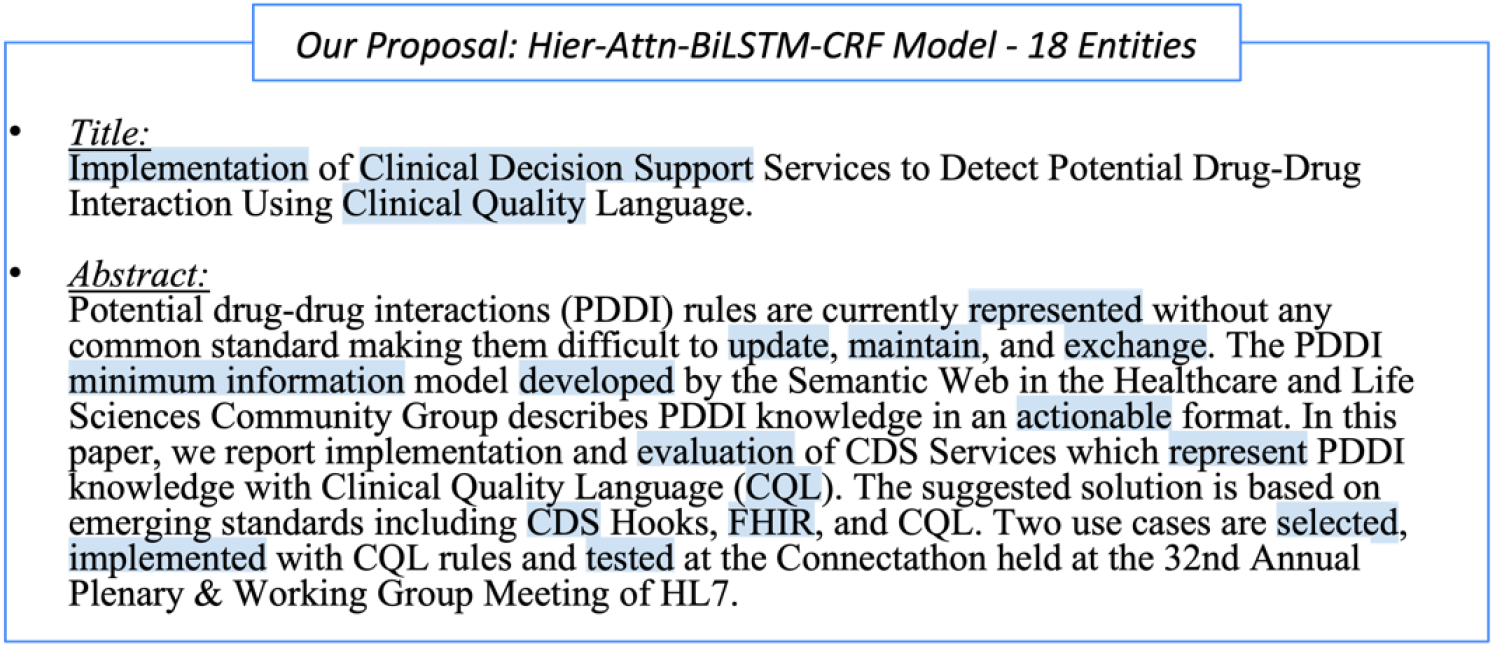
Entities identified in the sample CDSS article context with our Hier-Attn-BiLSTM-CRF model.

#### 3.4.1. Fine-tuning with Gold Standard (GS) labels

We fine-tuned [56] the Hier-Attn-BiLSTM-CRF model by mixing HDE and synthetic labeled documents in 0:100, 2:100, 4:100, 6:100, 8:100 proportions across 10 and 50 independent runs. Mixing 2–4 GS documents per 100 synthetic documents yielded the best trade-off, improving F1 from 55% to 84% and accuracy from 88% to 96% without exhausting HDE labels too quickly (Figure 8; Appendices E, F).

**Figure 8.**
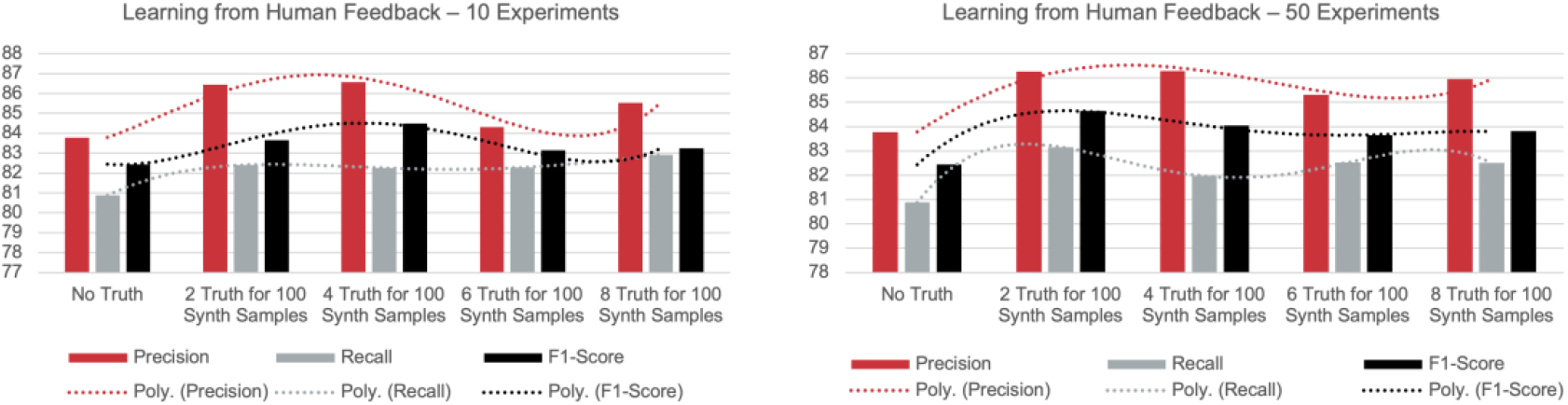
Results for fine-tuning with Gold Standard (GS) labels.

### 3.5. Additional experiments: robustness, baselines, and external validity

To assess robustness and external validity, we conducted additional experiments using 95% bootstrap confidence intervals (CIs; 2,000 resamples) on the same train/test split.

#### 3.5.1. Ablation and component attribution

A one-factor-at-a-time ablation (Appendix G, Table G1) shows that removing document-level attention, BiLM transfer, or pre-trained Word2Vec each changes GS91 F1 by less than ±0.6 points, whereas gold-standard fine-tuning is the dominant lever (+10.1 GS42, +3.9 GS91 F1). Additional controls (Appendix G, Table G2) confirm that the performance ceiling is set primarily by the label-generating process rather than any single architectural component.

#### 3.5.2. Fine-tuned transformer baselines

Three fine-tuned biomedical transformers (BioBERT, SciBERT, PubMedBERT) cluster tightly on GS42 (∼46 F1) with heavily overlapping CIs, only modestly ahead of the proposed model (∼44 F1). On GS91, the Hier-Attn-BiLSTM-CRF (61.3 F1) exceeds all transformers (∼54 F1) by ≈7 points—above every transformer CI—at a fraction of their ∼110M parameters (Appendix H, Table H1). Zero-shot controls (Appendix H, Table H2) confirm that transformer performance derives from task-specific fine-tuning (0–13 F1 without it).

#### 3.5.3. Train-Validation split sensitivity

An eight-point sweep of the train fraction (0.10–1.00) shows GS91 F1 increasing monotonically (55.05 → 62.70), confirming the original 1:2 split is a conservative lower bound (Appendix I, Table I1).

#### 3.5.4. Multi-seed variance

Training over five seeds yields SD ≈ 0.2–0.4 F1, confirming single-run reliability (Appendix J, Table J1). The Hier-Attn advantage on GS91 (+0.36 mean F1) is small but consistent.

#### 3.5.5. Error Analysis

Errors decompose into false positives (46.4%), false negatives (30.7%), boundary errors (22.8%), and nested-phrase overlaps (23.0%) (Appendix K, Tables K1–K2). The dominant failure mode is over-generation of generic spans inherited from synthetic labels. Approximately 11% of gold spans are structurally unmatchable under the tokenizer, identifying partial-match reporting as a high-value future improvement.

#### 3.5.6. Public corpus benchmarking

Models trained on the sparser KPBioMed and PubMedAKE corpora (ρ ≈ 0.06–0.08 vs. CDSS ρ ≈ 0.19–0.20) show healthy in-domain F1 (43.6–53.5) but collapsed cross-domain GS F1 (2.2–12.4) across all architectures, confirming a genuine domain gap and supporting the value of in-domain synthetic labels (Appendix L, Table L1).

## 4. Discussion

Limited HDE-labeled data is a common challenge in clinical NLP. Our work demonstrates that synthetic labels generated through domain adaptation, combined with hierarchical attentions along with the document-level context and minimal gold-standard fine-tuning, yield a practical KP identification pipeline. The ML model facilitates screening of candidate terms for CDSS ontology but does not replace expert judgment.

### 4.1. Results interpretation

KP identification is more complex than binary tagging—HDE draws on deep domain knowledge to judge relevance. Domain adaptation of sciSpacy produced substantially better synthetic labels than the base model, with a 2-fold F1 improvement on GS133 (Appendix D, Table D1). The hierarchical attention mechanism improved accuracy from 78% to 88% by providing document-level context to each word representation (Figure 6). Fine-tuning with GS labels in 2:100–4:100 HDE-to-synthetic label ratios further improved F1 from 55% to 84% (Figure 8), demonstrating the value of efficient use of scarce expert labels.

We used Word2Vec for pre-training due to its straightforward embedding matrix transfer and employed a dynamic batch loader (one document per batch) to maintain document-level context—increasing training time 2–3× but yielding clear accuracy gains. Visual inspection (Figure 7) shows identified KPs closely approximate GS labels, though the strict exact-match metric [48, 57] penalizes partial overlaps.

### 4.2. Robustness, baselines, and external validity

The additional experiments (Section 4.5) confirm that gold-standard fine-tuning is the dominant performance lever, while individual architectural components contribute small, within-noise effects. Against fine-tuned transformers, the Hier-Attn-BiLSTM-CRF is competitive (GS42) and sometimes favorable (for GS91, ≈7 F1 margin). The split-sensitivity sweep, multi-seed analysis, and public-corpus benchmarking together confirm stability, absence of split bias, and a genuine domain gap that validates the in-domain synthetic-label approach.

### 4.3. Challenges

Generating HDE labels is an expert-intensive process. Random sampling for annotation mitigates but does not guarantee topical diversity. The same selection-bias risk applies to the fine-tuning data loader, which currently selects documents without diversification.

### 4.4. Significance of the work

The contribution of this study lies in integrating established components—CDSS keyphrase identification task formulation, domain-adapted synthetic labels, document-level attention, and minimal gold-standard fine-tuning—into a practical, resource-efficient pipeline. This is a feasibility study with competitive and sometimes favorable performance to contemporary transformer baselines for our particular task. The approach is particularly relevant for automating curation of domain-specific ontologies under annotation-scarce conditions.

### 4.5. Limitations

Several limitations remain. First, some low-data control experiments (Appendix G, Table G2) used a single seed; multi-seed confidence intervals would strengthen directional claims. Second, the strict exact-match metric and tokenizer mismatches leave ≈11% of gold standards structurally unmatchable, understating practical usefulness; relaxed/partial-match reporting is a concrete next step. Third, this work predates the current LLM era; comparing LLM prompting/fine-tuning against our model remains valuable future work. Finally, a full downstream evaluation—measuring ontology coverage gain, curation-time reduction, and rule-component identification— was not performed and could be the primary next step.

## 5. Conclusion

This feasibility study presents a practical, resource-efficient framework for CDSS keyphrase identification under limited HDE annotation. The Hier-Attn-BiLSTM-CRF achieves competitive and sometimes favorable performance with fine-tuned biomedical transformers, while the dominant performance lever is the gold-standard fine-tuning step rather than any single architectural component. Multi-seed, split-sensitivity, and public-corpus experiments confirm result stability and a genuine CDSS domain gap. A full downstream evaluation of the role of the framework for CDSS ontology construction is the primary next step.

## Data Availability

The code is available on GitHub: https://github.com/xjing16/cdss4pcp_nlpml_pipeline.

## Abbreviations

NLP: Natural language processing
CDSS: Clinical decision support system
HDE: Human domain expert
BiLSTM: Bidirectional long short-term memory
BiLM: Bidirectional language model
CRF: Conditional random field
GS: Gold standard
KP: Keyphrase

## 6. Declarations

### Ethics approval and consent to participate

Not applicable to our study. We used publicly available publications in this study.

### Consent for publication

All co-authors read and approved the manuscript for publication.

### Availability of data and materials

The codes used in this study are available via GitHub:

https://github.com/xjing16/cdss4pcp_nlpml_pipeline

### Authors’ contributions

RG coded, established the initial pipeline, conducted the initial experiments, wrote the first draft of the manuscript, and revised it; KK and SA continued improving the pipeline, conducted more comprehensive experiments, and significantly revised the manuscript; NH and LR provided valuable input at different stages of the experiments; XJ provided significant feedback in designing, conducting, and revising the study and the manuscript iteratively; all other co-authors participated in the design and revision of the study and the manuscript.

## Acknowledgements

We acknowledge Clemson University for the generous allotment of compute time on the Palmetto cluster for our experimentation, and we thank Professor Feng Luo for sharing the computing resources. Also, we thank the anonymous reviewers for their detailed and insightful comments on earlier drafts of this paper.

## References

[1] Marco-Ruiz L, Bellika JG. Semantic Interoperability in Clinical Decision Support Systems: A Systematic Review. Stud Health Technol Inform 2015;216:958.

[2] Fernández-Breis JT, Vivancos-Vicente PJ, Menárguez-Tortosa M, et al. Using semantic technologies to promote interoperability between electronic healthcare records’ information models. Conf Proc IEEE Eng Med Biol Soc 2006;2006:2614–7. doi: 10.1109/iembs.2006.259686.

[3] Lobach D, Sanders G, Bright T, et al. Enabling Health Care Decision making Through Clinical Decision Support and Knowledge Management. Evidence Report No. 203. (Prepared by the Duke Evidence-based Practice Center under Contract No. 290-2007-10066-I.) AHRQ Publication No. 12-E001-EF.; 2012; Rockville, MD.

[4] Jing X, Himawan L, Law T. Availability and usage of clinical decision support systems (CDSS) in office-based primary care settings in the United States of America. BMJ Health & Care Informatics (under revision) 2019. 10.1136/bmjhci-2019-100015

[5] Gruber T. What is an ontology? Knowledge Acquisition 1993;5:199–220.

[6] Rector A. Foundations of the Semantic Web: Ontology Engineering. 2005;2006

[7] Jing X, Min H, Gong Y, et al. A systematic review of ontology-based clinical decision support system rules: usage, management, and interoperability. medRxiv; 2022. 10.1101/2022.05.11.22274984.

[8] He Zhiyong, Wang Zanbo, Wei Wei, Feng Shanshan, Mao Xianling, Jiang Sheng. (2020). A Survey on Recent Advances in Sequence Labeling from Deep Learning Models. 10.48550/arXiv.2011.06727.

[9] Kazi Saidul Hasan, Vincent Ng. 2014. Automatic Keyphrase Extraction: A Survey of the State of the Art. In Proceedings of the 52nd Annual Meeting of the Association for Computational Linguistics (Volume 1: Long Papers), pages 1262–1273, Baltimore, Maryland. Association for Computational Linguistics. 10.3115/v1/P14-1119.

[10] Salton, G; McGill, M. J. (1986). Introduction to modern information retrieval. McGrawHill. ISBN 978-0-07-054484-0. 10.5555/576628

[11] Hasan K. S., Ng V. (2010). Conundrums in unsupervised keyphrase extraction: Making sense of the state-of-the-art. In: Proceedings of the 23rd International Conference on Computational Linguistics, Beijing, China, pp. 365–373. https://dl.acm.org/doi/proceedings/10.5555/1944566.

[12] Amati, G. (2009). BM25. In: Liu, L., Ozsu, M.T. (eds) Encyclopedia of Database Systems. Springer, Boston, MA. 10.1007/978-0-387-39940-9921

[13] J. Berkhout, “Google’s PageRank algorithm for ranking nodes in general networks,”2016 13th International Workshop on Discrete Event Systems (WODES), Xi’an, China, 2016, pp. 153–158, doi: 10.1109/WODES.2016.7497841.

[14] Florian Boudin. 2018. Unsupervised Keyphrase Extraction with Multipartite Graphs. In Proceedings of the 2018 Conference of the North American Chapter of the Association for Computational Linguistics: Human Language Technologies, Volume 2 (Short Papers), pages 667–672, New Orleans, Louisiana. Association for Computational Linguistics. 10.48550/arXiv.1803.08721

[15] Corina Florescu and Cornelia Caragea. 2017. PositionRank: An Unsupervised Approach to Keyphrase Extraction from Scholarly Documents. In Proceedings of the 55th Annual Meeting of the Association for Computational Linguistics (Volume 1: Long Papers), pages 1105–1115, Vancouver, Canada. Association for Computational Linguistics. 10.18653/v1/P17-1102

[16] Adrien Bougouin, Florian Boudin, and Béatrice Daille. 2013. TopicRank: Graph-Based Topic Ranking for Keyphrase Extraction. In Proceedings of the Sixth International Joint Conference on Natural Language Processing, pages 543–551, Nagoya, Japan. Asian Federation of Natural Language Processing.

[17] Raschka, S. (2014). Naive Bayes and Text Classification I -Introduction and Theory. ArXiv, abs/1410.5329.

[18] Quinlan, J.R. Induction of decisiontrees. Mach Learn 1, 81–106 (1986). 10.1007/BF00116251

[19] Evgeniou, Theodoros & Pontil, Massimiliano. (2001). Support Vector Machines: Theory and Applications. 2049. 249–257. 10.1007/3-540-44673-712.

[20] Huang, Z.H., Xu, W., Yu, K.: Bidirectional LSTM-CRF models for sequence tagging. CoRR, abs/1508.01991 (2015)

[21] Jiang Y, Zhao T, Chai Y, et al. (2020) Bidirectional LSTM-CRF models for keyword extraction in Chinese sport news. In: MIPPR 2019: Pattern Recognition and Computer Vision. International Society for Optics and Photonics, 11430: 114300H. 10.48550/arXiv.1508.01991

[22] J. Li, A. Sun, J. Han and C. Li,” A Survey on Deep Learning for Named Entity Recognition,” in IEEE Transactions on Knowledge and Data Engineering, vol. 34, no. 1, pp. 50–70, 1 Jan. 2022, doi: 10.1109/TKDE.2020.2981314. https://doi.org/10.1109/TKDE.2020.2981314

[23] Nikolaos Giarelis and Nikos Karacapilidis. 2024. Deep learning and embeddings-based approaches for keyphrase extraction: a literature review. Knowl. Inf. Syst. 66 (11): 6493–6526 (2024). 10.1007/s10115-024-02164-w

[24] Muhammad Umair, Tangina Sultana, Young-Koo Lee. Pre-trained language models for keyphrase prediction: A review. ICT Express. 10 (4):871–890 (2024.). 10.1016/j.icte.2024.05.015.

[25] M. B. Lakshmi, M. D. Padmanabhuni, B. S. P. Katragadda, D. Nakkina and S. J. S. Pusuluri, “Keyword Identification and Summarization in a Text by Using Deep Learning Based Models,”2025 International Conference on Inventive Computation Technologies (ICICT), Kirtipur, Nepal, 2025, pp. 1307–1313, doi: 10.1109/ICICT64420.2025.11005054.

[26] Zeyer Albert, Bahar Parnia, Irie Kazuki, Schluter Ralf, Ney Hermann. (2019). A Comparison of Transformer and LSTM Encoder Decoder Models for ASR. 8–15. 10.1109/ASRU46091.2019.9004025.

[27] Imran Ahamad Sheikh, Emmanuel Vincent, Irina Illina. Transformer versus LSTM Language Models Trained on Uncertain ASR Hypotheses in Limited Data Scenarios. LREC 2022 -13th Language Resources and Evaluation Conference, Jun 2022, Marseille, France. hal-03362828v2

[28] Hochreiter S., Schmidhuber J”urgen. (1997). Long short-term memory. Neural Computation, 9(8), 1735–1780. 10.1162/neco.1997.9.8.1735

[29] Lafferty J. D., McCallum A., Pereira F. C. N. (2001). Conditional Random Fields: Probabilistic Models for Segmenting and Labeling Sequence Data. Proceedings of the Eighteenth International Conference on Machine Learning (p./pp. 282–289), San Francisco, CA, USA: Morgan Kaufmann Publishers Inc. ISBN: 1-55860-778-1. https://dl.acm.org/doi/proceedings/10.5555/645530

[30] Sahrawat, D. et al. (2020). Keyphrase Extraction as Sequence Labeling Using Contextualized Embeddings. In:, et al. Advances in Information Retrieval. ECIR 2020. Lecture Notes in Computer Science (), vol 12036. Springer, Cham. 10.1007/978-3030-45442-541.

[31] Zichao Yang, Diyi Yang, Chris Dyer, Xiaodong He, Alex Smola, and Eduard Hovy. 2016. Hierarchical Attention Networks for Document Classification. In Proceedings of the 2016 Conference of the North American Chapter of the Association for Computational Linguistics: Human Language Technologies, pages 1480–1489, San Diego, California. Association for Computational Linguistics. 10.18653/v1/N16-1174

[32] Xu, Guohai & Wang, Chengyu & He, Xiaofeng. (2018). Improving Clinical Named Entity Recognition with Global Neural Attention: Second International Joint Conference, APWeb-WAIM 2018, Macau, China, July 23–25, 2018, Proceedings, Part II. 10.1007/9783-319-96893-320.

[33] Mikhailov, Vladislav & Shavrina, Tatiana. (2020). Domain-Transferable Method for Named Entity Recognition Task. 83–92. https://doi.org/10.5121/csit.2020.101407. 10.48550/arXiv.2011.12170

[34] Kulkarni, Vivek & Mehdad, Yashar & Chevalier, Troy. (2016). Domain Adaptation for Named Entity Recognition in Online Media with Word Embeddings. 10.48550/arXiv.1612.00148

[35] Neumann, M., King, D., Beltagy, I., & Ammar, W. (2019). ScispaCy: Fast and Robust Models for Biomedical Natural Language Processing. ArXiv, abs/1902.07669.

[36] Honnibal, M., & Montani, I. (2017). spaCy 2: Natural language understanding with Bloom embeddings, convolutional neural networks and incremental parsing.

[37] Yoshua Bengio, Réjean Ducharme, Pascal Vincent, and Christian Janvin. 2003. A neural probabilistic language model. J. Mach. Learn. Res. 3, null (3/1/2003), 1137–1155. https://dl.acm.org/toc/jmlr/2003/3/null

[38] L. Rabiner and B. Juang,” An introduction to hidden Markov models,” in IEEE ASSP Magazine,vol. 3, no. 1, pp. 4–16, Jan 1986, doi: 10.1109/MASSP.1986.1165342.

[39] Sachan, D.S., Xie, P., Sachan, M., & Xing, E.P. (2017). Effective Use of Bidirectional Language Modeling for Transfer Learning in Biomedical Named Entity Recognition. Machine Learning in Health Care. 10.48550/arXiv.1711.07908

[40] Lev Ratinov and Dan Roth. 2009. Design Challenges and Misconceptions in Named Entity Recognition. In Proceedings of the Thirteenth Conference on Computational Natural Language Learning (CoNLL-2009), pages 147–155, Boulder, Colorado. Association for Computational Linguistics. https://dl.acm.org/doi/proceedings/10.5555/1596374

[41] Saad, F., Aras, H., Hackl-Sommer, R. (2020). Improving Named Entity Recognition for Biomedical and Patent Data Using Bi-LSTM Deep Neural Network Models. In: Métais, E., Meziane, F., Horacek, H., Cimiano, P. (eds) Natural Language Processing and Information Systems. NLDB 2020. Lecture Notes in Computer Science, vol 12089. Springer, Cham. 10.1007/978-3-030-51310-83

[42] Mikolov, Tomas et al. “Efficient Estimation of Word Representations in Vector Space.” International Conference on Learning Representations (2013). 10.48550/arXiv.1301.3781

[43] M. Schuster and K. K. Paliwal,” Bidirectional recurrent neural networks,” in IEEE Transactions on Signal Processing, vol. 45, no. 11, pp. 2673–2681, Nov. 1997, doi: 10.1109/78.650093.

[44] Luo L, Yang Z, Yang P, Zhang Y, Wang L, Lin H, Wang J. An attention-based BiLSTMCRF approach to document-level chemical named entity recognition. Bioinformatics. 2018 Apr 15;34(8):1381–1388. doi: 10.1093/bioinformatics/btx761. PMID: 29186323.

[45] Devlin, J., Chang, M.-W., Lee, K. & Toutanova, K. (2018). BERT: Pre-training of Deep Bidirectional Transformers for Language Understanding 10.48550/arXiv.1810.04805.

[46] Sainbayar Sukhbaatar, Arthur Szlam, Jason Weston, and Rob Fergus. 2015. End-to-end memory networks. arXiv preprint arXiv:1503.08895.

[47] A. Viterbi,” Error bounds for convolutional codes and an asymptotically optimum decoding algorithm,” in IEEE Transactions on Information Theory, vol. 13, no. 2, pp. 260–269, April 1967, doi: 10.1109/TIT.1967.1054010.

[48] Hiroki Nakayama.” A Python framework for sequence labeling evaluation” 2018. https://github.com/chakki-works/seqeval

[49] Jing X, Indani A, Hubig NC, et al. A systematic approach to configuring MetaMap for optimal performance. Methods Inf Med 2022 doi: 10.1055/a-1862-0421

[50] Maël Houbre, Florian Boudin, and Beatrice Daille. 2022. A Large-Scale Dataset for Biomedical Keyphrase Generation. In Proceedings of the 13th International Workshop on Health Text Mining and Information Analysis (LOUHI), pages 47–53, Abu Dhabi, United Arab Emirates (Hybrid). Association for Computational Linguistics.

[51] Jiasheng Sheng, Zelalem Gero, and Joyce C. Ho. 2022. PubMed Author-assigned Keyword Extraction (PubMedAKE) Benchmark. In Proceedings of the 31st ACM International Conference on Information & Knowledge Management (CIKM ‘22). Association for Computing Machinery, New York, NY, USA, 4470–4474. 10.1145/3511808.355767

[52] Collobert, R., Weston, J., Bottou, L., Karlen, M., Kavukcuoglu, K., Kuksa, P.: Natural language processing (almost) from scratch. J. Mach. Learn. Res. 12, 2493–2537 (2011). 10.48550/arXiv.1103.0398

[53] Jinhyuk Lee, Wonjin Yoon, Sungdong Kim, Donghyeon Kim, Sunkyu Kim, Chan Ho So, and Jaewoo Kang. 2020. BioBERT: a pre-trained biomedical language representation model for biomedical text mining. Bioinformatics, 36(4):1234–1240. 10.1093/bioinformatics/btz682

[54] Yu Gu, Robert Tinn, Hao Cheng, Michael Lucas, Naoto Usuyama, Xiaodong Liu, Tristan Naumann, Jianfeng Gao, and Hoifung Poon. 2021. Domain-Specific Language Model Pretraining for Biomedical Natural Language Processing. ACM Trans. Comput. Healthcare 3, 1, Article 2 (January 2022), 23 pages. 10.1145/3458754

[55] Iz Beltagy, Kyle Lo, and Arman Cohan. 2019. SciBERT: A Pretrained Language Model for Scientific Text. In Proceedings of the 2019 Conference on Empirical Methods in Natural Language Processing (EMNLP). arXiv:1903.10676.

[56] Syed, Muzamil Hussain, and Sun-Tae Chung. 2021.” MenuNER: Domain-Adapted BERT Based NER Approach for a Domain with Limited Dataset and Its Application to Food Menu Domain” Applied Sciences 11, no. 13: 6007. 10.3390/app11136007

[57] Nancy Chinchor and Beth Sundheim. 1993. MUC-5 Evaluation Metrics. In Fifth Message Understanding Conference (MUC-5): Proceedings of a Conference Held in Baltimore, Maryland, August 25–27, 1993.

