## Appendices A to I for "Keyphrase Identification Using Minimal Labeled Data with Hierarchical Contexts and Transfer Learning"

#### 8. Appendices

### Appendix A. Detailed Encoder-Decoder Diagram

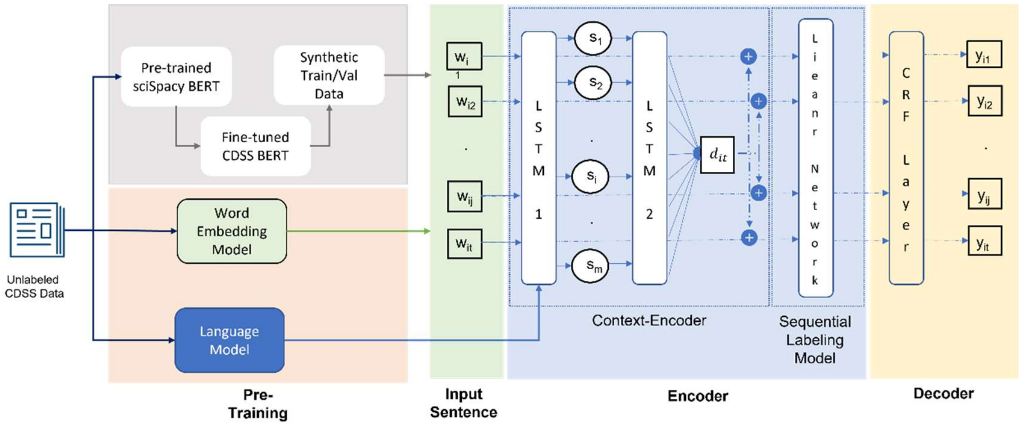


### Appendix B. Dataset Details

**Table B1.** Showing explicit details of the CDSS dataset during preprocessing

| Type | Abstracts Number |
|---|---|
| Total after parsing PubMed XML | 3326 |
| HDE-labeled Set 1 (GS42) | 42 |
| ACM abstracts [8]<br>+<br>HDE-labeled Set 2 (PMIDs not in XML) [4] | 8+4 = 12 |
| New total with duplicates<br>(Some articles from GS42 are in full text XML) | 3380 |
| Abstracts (<3 sentences ~little/no abstract) | 99 |
| New total with duplicates<br>(After removing abstracts with <3 sentences) | 3281<br>(1093 train + 2188 test) |
| HDE-labeled Set 2 (GS91)<br>(ACM 8 + PubMed 83) | 83 + 8 = 91 |
| Total GS | 91 + 42 = 133 |
| Final total<br>(Synthetic-labeled dataset)<br>(After removing GS 133 from full dataset) | 3148<br>(1049 train + 2099 test) |

The code is available on GitHub: https://github.com/xjing16/cdss4pcp_nlpml_pipeline.

#### Appendix C. Entities identified on sample CDSS abstract

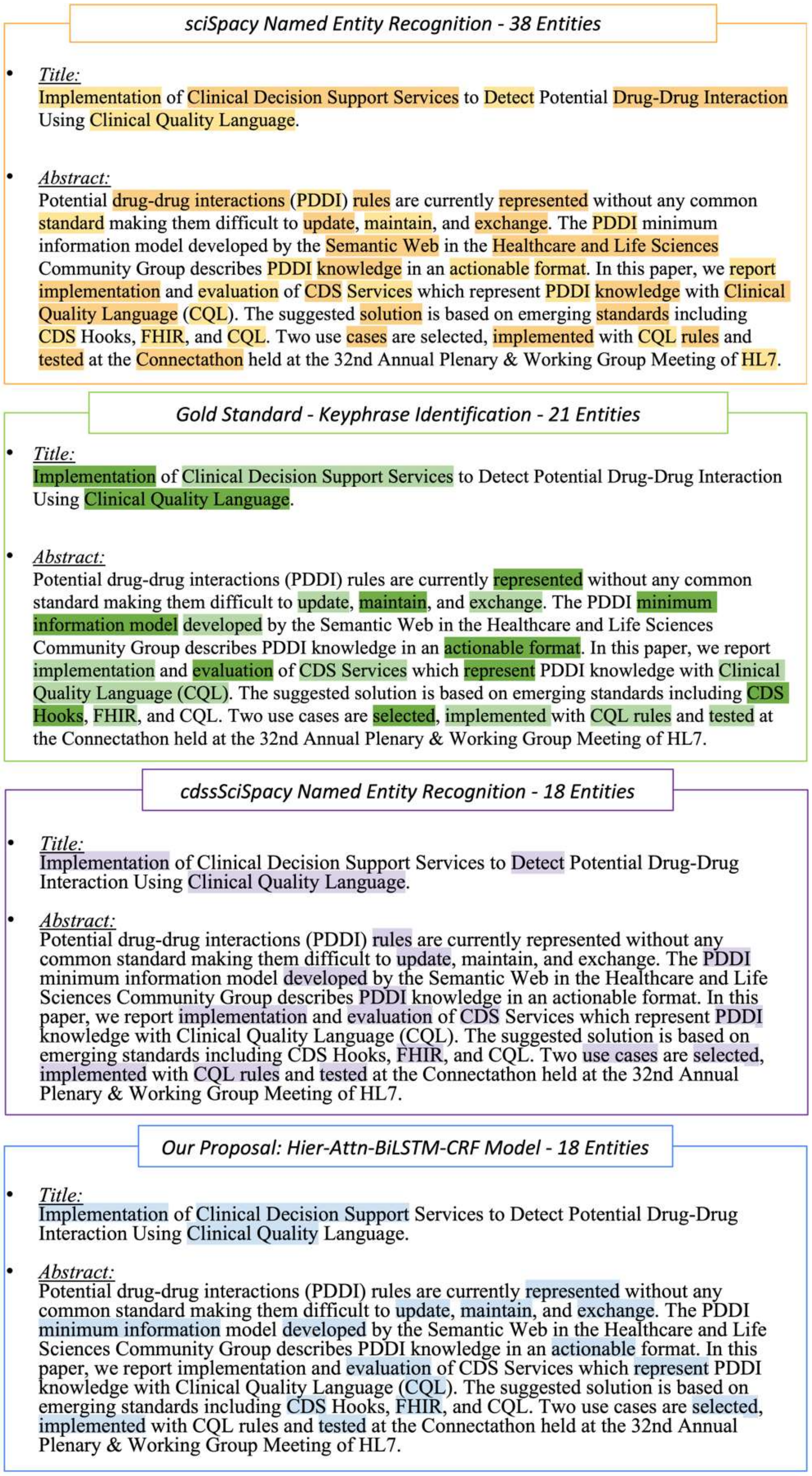


**Figure C1.** Entities identified on sample CDSS abstract demonstrate differences between human-labeled Gold Standards and different models (sciSpacy, cdssSciSpacy, and Hier-Attn-BiLSTM-CRF).

### Appendix D. Evaluation of fine-tuning sciSpacy model for CDSS

**Table D1.** Evaluation of fine-tuning sciSpacy model for CDSS (scale 0-1)

| Fine-Tune | Base | Model | Train Dataset | GS Dataset | Precision | Recall | Accuracy | F1-Score |
|---|---|---|---|---|---|---|---|---|
| Level 0 | sciSpacy | sciSpacy (en_core_sci_lg) | 3281 from PubMed | 42 | 0.61 | 0.18 | 0.93 | 0.27 |
| | | | | 91 | 0.59 | 0.23 | 0.97 | 0.33 |
| | | | | 133 | 0.62 | 0.22 | 0.96 | 0.33 |
| **Level 1** | **sciSpacy** | **cdssSciSpacy** | Synthetic CDSS (1866 Train / 622 Val) | 42 | 0.70 | 0.38 | 0.97 | 0.5 |
| | | | | 91 | 0.73 | **0.64** | **0.99** | **0.68** |
| | | | | 133 | **0.74** | 0.59 | **0.99** | 0.66 |
| Level 2 | cdssSciSpacy | cdssSciSpacy GS42 | 42 GS (33 Train / 9 Val) | 91 | 0.57 | **0.64** | **0.99** | 0.60 |
| Level 2 | cdssSciSpacy | cdssSciSpacy GS91 | 91 GS (72 Train / 19 Val) | 42 | 0.66 | 0.38 | 0.97 | 0.48 |
| Level 2 | sciSpacy | sciSpacy GS42 | 42 GS (33 Train / 9 Val) | 91 | 0.57 | 0.54 | **0.99** | 0.55 |
| Level 2 | cdssSciSpacy | cdssSciSpacy GS66 [1] | 66 GS (52 Train / 14 Val) | 67 | 0.63 | 0.62 | **0.99** | 0.62 |

[1]Repeated experiment 50 times on random samples of GS 133.

### Appendix E. Metrics for fine-tuning on GS

**Table E1.** Fine-tuning with GS labels - 10 experiments

| GS | 0 | 2 | 4 | 6 | 8 | 10 | 12 |
|---|---|---|---|---|---|---|---|
| Precision | 83.78 ± 11.12 | 86.43 ± 5.78 | 86.56 ± 9.86 | 84.32 ± 4.99 | 85.54 ± 6.94 | 84.35 ± 9.92 | 85.99 ± 10.75 |
| Recall | 80.88 ± 4.89 | 82.41 ± 7.85 | 82.26 ± 5.69 | 82.28 ± 2.21 | 82.91 ± 9.41 | 81.80 ± 15.32 | 82.99 ± 7.20 |
| Accuracy | 95.62 ± 0.93 | 96.21 ± 0.51 | 96.24 ± 1.04 | 95.65 ± 0.58 | 95.73 ± 1.12 | 95.88 ± 0.56 | 96.13 ± 0.80 |
| F1-Score | 82.44 ± 4.82 | 83.66 ± 3.79 | 84.48 ± 5.68 | 83.14 ± 2.05 | 83.35 ± 7.21 | 82.81 ± 4.38 | 84.22 ± 7.42 |

**Table E2.** Fine-tuning with GS labels - 50 experiments

| GS | 0 | 2 | 4 | 6 | 8 | 10 | 12 |
|---|---|---|---|---|---|---|---|
| Precision | 83.78 ± 10.21 | 86.27 ± 8.28 | 86.29 ± 9.15 | 85.30 ± 8.01 | 85.95 ± 9.16 | 85.91 ± 8.70 | 86.22 ± 11.35 |
| Recall | 80.88 ± 4.49 | 83.16 ± 8.14 | 81.97 ± 11.91 | 82.53 ± 6.66 | 82.50 ± 7.83 | 82.45 ± 11.26 | 82.85 ± 8.20 |
| Accuracy | 95.62 ± 0.85 | 96.24 ± 0.74 | 96.16 ± 1.04 | 95.92 ± 0.72 | 96.06 ± 0.82 | 96.08 ± 0.87 | 96.27 ± 0.85 |
| F1-Score | 82.44 ± 4.43 | 84.65 ± 5.42 | 84.05 ± 7.36 | 83.65 ± 4.54 | 83.81 ± 6.05 | 83.91 ± 6.61 | 84.43 ± 6.59 |

### Appendix F. Plots for fine-tuning on GS

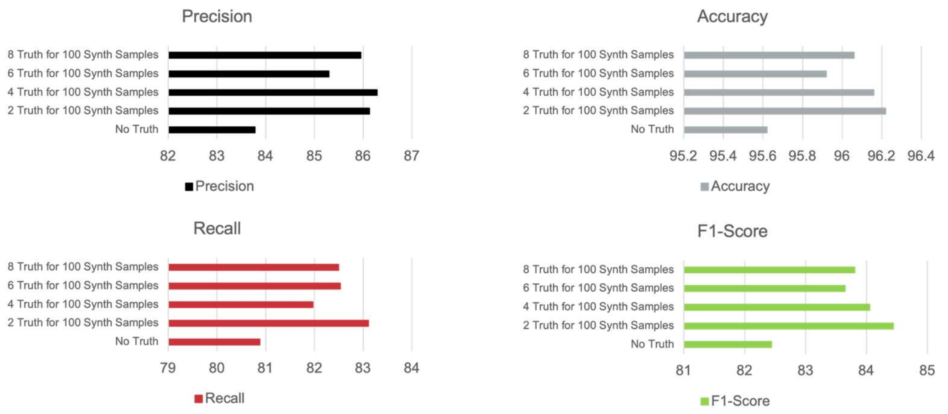


**Figure F1.** Plot evaluation metrics for fine-tuning with GS labels - 50 experiments

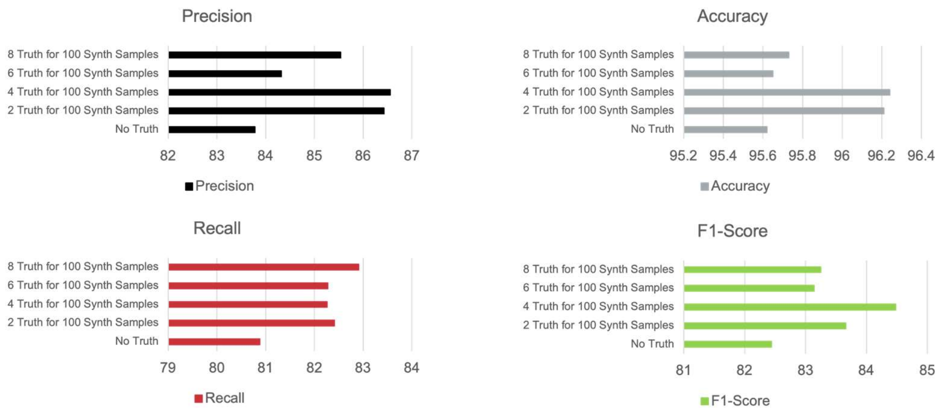


**Figure F2.** Plot evaluation metrics for fine-tuning with GS labels - 10 experiments

### Appendix G. Ablation and component-attribution study

All values are strict entity-level SeqEval micro-F1 (%) on the held-out gold standards, using the tuned configuration of the proposed model (IOBES tagging, AdamW, gated document-level attention, 20 epochs), seed 42 unless noted.

**Table G1.** One-factor-at-a-time ablation against the full model

| Configuration | Change vs. full model | GS42 F1 | GS91 F1 | Effect on GS91 |
|---|---|---|---|---|
| Full Model | - | 43.99 | 61.13 | - |
| - document attention | Remove sent/doc attention | 44.53 | 61.50 | +0.37 |
| - BiLM Transfer | Remove pre-trained BiLM | 44.21 | 60.93 | -0.20 |
| - pre-trained Word2Vec | Random embeddings | 44.46 | 61.32 | +0.19 |
| + gold standard fine-tuning (10 epochs) | Add short GS fine tuning | 54.12 | 65.02 | +3.89 |

**Table G2.** Low-data control experiments isolating the source of generalization (effect on GS91 F1)

| Manipulation | Setting | Effect on GS91 F1 |
|---|---|---|
| Untrained Baseline | 0 epochs, random CRF head | 13 (vs. ~55) |
| Remove BiLM | Remove BiLM, Keep Word2Vec | <0.5 (sign flips across fractions) |
| Remove all pre-training | Fully random init, trainable embeddings | ≈ 0 (within ±1.8) |
| Freeze embeddings | frozen Word2Vec vs. frozen random | < 2.7 (mean ≈ 0) |
| Remove hand-crafted features | drop POS / WFOF / LEN / TI / TR | < 1 |
| Switch architecture (synthetic data) | 12 GNA configs (4 arch × 3 schemes) | all cluster within ≈3 F1 (51–53) |
| Switch architecture (real PubMedAKE) | GNA vs. domain transformers | transformers +6–7 over GNA |
| Synthetic → real author keyphrases | change label source | −16 to −20 (≈66 → ≈41–46) |

### Appendix H. Transformer baselines and zero-shot controls

**Table H1.** Fine-tuned biomedical transformers on the same split, with 95% document-bootstrap CIs (2,000 resamples; strict entity F1, %)

| Model | GS42 F1 | GS42 95% CI | GS91 F1 | GS91 95% CI |
|---|---|---|---|---|

| | | | | |
|---|---|---|---|---|
| PubMedBERT | 46.91 | [43.30, 50.70] | 54.33 | [51.44, 56.96] |
| BioBERT | 45.13 | [41.70, 48.64] | 53.65 | [50.73, 56.22] |
| sciBERT | 46.19 | [42.69, 49.77] | 54.26 | [51.39, 57.00] |
| Hier-Attn-BiLSTM-CRF | 44.17 | [41.80, 46.78] | 61.27 | [59.34, 63.08] |

**Table H2.** Low-data control experiments isolating the source of generalization (effect on GS91 F1)

| **Route** | Mode | Trained on KP Labels? | GS42 F1 | GS91 F1 |
|---|---|---|---|---|
| PubMed BERT, random head | A | No | 0.00 | 0.07 |
| BioBERT, random head | A | No | 1.62 | 1.16 |
| sciBERT, random head | A | No | 6.45 | 7.52 |
| BioMedical-NER head transfer | B | No | 11.58 | 13.27 |
| Fine-tuned transformers (Table H1) | C | Yes | 45.13-46.91 | 53.65-54.33 |

### Appendix I. Train-validation split sensitivity

**Table I1.** Full eight-point train-fraction sweep with the pre-trained language model held constant (strict entity F1, %)

| **Train fraction** | Train docs | GS42 F1 | GS91 F1 |
|---|---|---|---|
| 0.1 | 328 | 41.64 | 55.05 |
| 0.25 | 820 | 42.37 | 55.76 |
| 0.33 (original ≈1:2) | 1049 | 43.28 | 58.83 |
| 0.50 | 1640 | 44.10 | 59.82 |
| 0.70 | 2297 | 44.22 | 60.75 |
| 0.75 | 2461 | 43.52 | 60.80 |
| 0.80 | 2625 | 45.07 | 62.08 |
| 1.00 | 3280 | 44.42 | 62.70 |

### Appendix J. Multi-Seed Variance

**Table J1.** Mean ± SD over five random seeds (13, 42, 101, 202, 777; strict entity F1, %)

| **Model** | **GS42 F1 (mean ± SD)** | **GS91 F1 (mean ± SD)** |
|---|---|---|
| BiLSTM-CRF | 44.29 ± 0.39 | 61.13 ± 0.22 |
| Hier-Attn-BiLSTM-CRF | 44.52 ± 0.34 | 61.49 ± 0.24 |

### Appendix K. Error Taxonomy

**Table K1.** Fine-tuned transformers, GS91 (seed 42)

| Model | TP | FP | FN | Boundary | Nested | Strict P | Strict R |
|---|---|---|---|---|---|---|---|
| PubMedBERT | 1299 | 1096 (46.4%) | 726 (30.7%) | 539 (22.8%) | 543 (23.0%) | 44.27 | 50.66 |
| BioBERT | 1249 | 982 (42.8%) | 816 (35.5%) | 499 (21.7%) | 507 (22.1%) | 45.75 | 48.71 |
| sciBERT | 1329 | 1204 (49.4%) | 701 (28.7%) | 534 (21.9%) | 546 (22.4%) | 43.33 | 51.83 |

**Table K2.** Fine-tuned transformers, GS42 (seed 42)

| Model | TP | FP | FN | Boundary | Nested | Strict P | Strict R |
|---|---|---|---|---|---|---|---|
| PubMedBERT | 597 | 757 (56.8%) | 331 (24.8%) | 244 (18.3%) | 238 (17.9%) | 37.36 | 50.94 |
| BioBERT | 564 | 691 (53.2%) | 373 (28.7%) | 235 (18.1%) | 225 (17.3%) | 37.85 | 48.12 |
| sciBERT | 603 | 776 (57.7%) | 312 (23.2%) | 257 (19.1%) | 242 (18.0%) | 36.86 | 51.45 |

### Appendix L. Public-dataset transfer study

**Table L1.** Public-corpus → CDSS gold-standard transfer after correcting the early-stopping artifact (strict entity F1, %)

| Model | In-domain F1 | GS42 F1 | GS91 F1 | Transfer Gap |
|---|---|---|---|---|
| BiLSTM-CRF (BIO) | 43.59 | 3.20 | 7.13 | ≈ 38 |
| Hier-Attn-gated (BIO) | 45.16 | 2.21 | 5.67 | ≈ 41 |
| BioBERT | 47.08 | 8.55 | 10.45 | ≈ 38 |
| SciBERT | 49.85 | 6.10 | 8.41 | ≈ 43 |
| PubMedBERT | 53.52 | 11.72 | 12.41 | ≈ 41 |

The code is available on GitHub: https://github.com/xjing16/cdss4pcp_nlpml_pipeline.
